# Autonomic and Neural Evaluation of the Dose Duration Response for Non-invasive Transcutaneous Auricular Vagus Nerve Stimulation

**DOI:** 10.1101/2025.06.03.25328424

**Authors:** Danielle L. Taylor, Christopher T. Sege, Samantha L. LaPorta, Cameron M. Robins, Bashar W. Badran, Mark S. George, Lisa M. McTeague

**Affiliations:** Department of Psychiatry and Behavioral Neurosciences, Wayne State University, Detroit, MI; Department of Psychiatry and Behavioral Sciences, Medical University of South Carolina, Charleston, SC; Department of Clinical Psychology, University of Southern Mississippi, Hattiesburg, MS; Ralph H. Johnson VA Healthcare System, Charleston, SC

## Abstract

Transcutaneous auricular vagus nerve stimulation (taVNS) is a candidate non-invasive brain stimulation-based clinical tool for a variety of psychiatric disorders. Despite suggestion of promise, null effects and inconsistencies in reporting have also been accumulating, suggesting a need to evaluate stimulation parameters that lead to more replicable, meaningful effects. The goal of this study was to evaluate the effects of dose duration (15 to 75 active minutes) of taVNS across multiple modalities (autonomics and electrocortical activity) and processes (attentional engagement and executive function) in 28 healthy control participants. Results demonstrate that higher doses (60-75 minutes) of active taVNS were required to adaptively modulate heart rate variability during cognitive engagement – a dose significantly higher than those being administered across the literature. We also demonstrate that P300 in an oddball task did not modulate with increasing dose. These results indicate that taVNS may specifically modulate motivationally salient systems, though it is important to note that this could be due to a ceiling effect of P300 in the context of this healthy sample. To our knowledge this is the first ever dose duration finding study for taVNS and future investigations are needed to continue to narrow the parameter space and replicate these findings in a larger sample with psychiatric symptoms.

---

Transcutaneous auricular vagus nerve stimulation (taVNS) is a non-invasive brain stimulation candidate for use in basic and translational research and as a potential clinical tool for a variety of psychiatric disorders (e.g., George et al., 2007; George et al., 2000; George et al., 2008). Stimulation is accomplished via two surface electrodes applied to the ear which deliver pulsed stimulation to vagally innervated ear regions (e.g., tragus and cymba concha (Badran et al., 2019). Some of the advantages of taVNS include that it is cost effective, has few side effects, it is safe and can be easily implemented in clinical or research settings without immediate MD oversight (Badran et al., 2022; Badran et al., 2020; Kim et al., 2022).

Studies have demonstrated a variety of effects including modulation of transdiagnostic mechanisms, such as inflexible stress responses, heightened anxious arousal, and impaired cognitive function (Badran, Dowdle et al., 2018; Badran, Mithoefer et al., 2018; Deuchars et al., 2018; Jenkins et al., 2019; Warren et al., 2020). However, among the diverse benefits, there have also been accumulating null effects, as well as inconsistencies in reporting (Farmer et al., 2021). For example, stimulation parameters (pulse width, stimulation duration, intensity, frequency, and active/sham locations) number of doses, stimulation device, and blinding procedures often are not fully reported. This contributes to null effects and replication difficulties. Null effects may also be attributable to a lack of consensus on the optimal stimulation parameters for taVNS, though some investigations have begun working to narrow the parameter space (Badran et al., 2019; Badran, Dowdle et al., 2018; Kreisberg et al., 2021). Prior to implementation of therapeutic taVNS or continued study of the biological or psychiatric effects, studies should systematically determine the optimal dose duration, which has not yet been determined. This was, therefore, the goal of the current study, to examine the effect of dose duration for taVNS across multiple modalities (autonomics and electrocortical activity) and processes (attentional engagement and executive function).

The vagus nerve is the longest cranial nerve containing a variety of fibers which maintain afferent parasympathetic control over the heart, lungs, and digestive track, while efferent fibers provide the prefrontal cortex with somatic and visceral feedback (Breit et al., 2018; Porges, 1985; Yuan & Silberstein, 2016). Vagal function is often indexed using vagal tone, such that cardiac activity is regulated by inhibitory vagal control (Tulppo et al., 2001). On the other hand, vagal innervation of cardiac responses has been modulated using implanted vagus nerve stimulation (VNS), resulting in increased parasympathetic activity (Zamotrinsky et al., 2001). These and other data are cited for evidence of the peripheral modulatory effects of VNS. Functional magnetic imaging (fMRI) data show increased connectivity of locus coeruleus-hippocampus tracts, evidencing the central modulatory effects of VNS (Berger et al., 2024). Whereas VNS has been FDA approved for a variety of conditions (e.g., epilepsy, treatment resistant depression, post-stroke motor rehabilitation), its use for outpatient psychiatric treatment and translational research is limited, as the VNS pulse generator must be surgically implanted in the chest cavity (Andalib et al., 2023; Austelle et al., 2024; George et al., 2008; Nahas et al., 2005; Sackeim et al., 2001). Therefore, a variant that is non-invasive and which theoretically activates comparable afferents and efferents, taVNS, has been increasingly investigated.

Studies have demonstrated some efficacy for taVNS across a variety of psychiatric symptoms including low mood, repetitive negative thinking (RNT), sleep disturbances, and anxiety (Badran et al., 2022; George et al., 2008; Kong et al., 2018; Szeska et al., 2020; Warren et al., 2019; Q. Zhang et al., 2024; Y. Zhang et al., 2024). Mechanistic findings (i.e., effects on autonomic and cognitive function), however, have been mixed. For example, several studies have demonstrated that taVNS decreases heart rate (HR) and increases vagal tone under certain conditions and parameters (Austelle et al., 2023; Badran, Mithoefer et al., 2018; Forte et al., 2022). On the other hand, investigations of indices of locus coeruleus norepinephrine (LC-NE) activation using the P300 event-related potential (ERP, an indicator of stimulus discrimination), eye tracking, and salivary alpha amylase as indices have been mixed, with some data supporting taVNS related adaptive modulation and others showing no effect (Burger et al., 2020; Warren et al., 2019; Warren et al., 2020).

The range in demonstrable effects may be attributable to high variability in the taVNS parameter space. Kim and colleagues, while conducting a meta-analysis of the safety profile of taVNS, present a range of durations between 1.5 and 360 minutes, whereas the majority of studies administer 20-30 minutes of stimulation (2022). However, there is no clear rationale for this more limited range of duration, nor has duration been systematically investigated. On the other hand, other parameters have been explored. For example, computational modeling has been utilized to investigate optimal electrode montages for active taVNS (tragus and cymba concha) with the highest likelihood of stimulating the auricular branch of the vagus nerve (ABVN), and control (ear lobe) with minimal ABVN innervation (Badran, Brown et al., 2018; Kreisberg et al., 2021). Additionally, efforts towards parameterization have been sought for pulse width, stimulation frequency, and train duration (Badran, Dowdle et al., 2018; Badran, Mithoefer et al., 2018). Evidenced by lower heart rate, 25 Hz and 500 μs pulse width, showed the largest reduction and 60 second on/off trains show neurophysiologic effects.

Therefore, the current investigation employed these stimulation parameters, while extending the systematic evaluation of the taVNS parameter space. Here, we evaluate multi-modal outputs for stimulation session duration to determine the dose response curve for taVNS in a sample of healthy participants. We hypothesized that greater dose durations than are typically cited in the literature (e.g., 20-30 mins) would be required to adaptively modulate neurophysiological outcomes. This hypothesis was made given FDA approved protocols administer implanted VNS for 30 minutes in depression and 60-90 minutes in stroke motor rehabilitation. TaVNS targets the vagus less directly through purported stimulation of the ABVN and is a less potent intervention than VNS, so it should therefore require greater duration for adaptive effects.

## Materials and Methods

### Overview

The current study aimed to determine the dose-duration response for taVNS on multiple vagally-mediated physiological measures in a blinded sham-controlled design. Twenty-eight healthy participants received 75 minutes of stimulation and were randomized to 5 different doses of active taVNS (15-75 minutes). Before and at the end of the stimulation period, participants completed a series of cognitive tasks while electroencephalography (EEG), heart rate, and skin conductance were collected. The study was approved by the institutional review board. All participants were provided informed consent before study participation. This paper represents preliminary findings, and full data are in preparation for peer review.

### Participants and Inclusion Criteria

Participants included 28 healthy community members, who were enrolled in the study after meeting the following study inclusion criteria 1) age 18 to 65 years; 2) English speaking 3) non-treatment seeking; 4) no diagnosis of COVID-19 in the past 14 days. Exclusion criteria included 1) facial or ear pain or recent ear trauma; 2) current psychiatric, seizure/anticonvulsant, blood pressure, antiarrhythmic, or beta-blocker medications, or medications that list seizures as a potential side effect; 3) implanted devices in the heart, neck, or head, or history of brain stimulation or brain surgery; 4) frequent or severe headaches; 5) history of seizures and/or epilepsy; 6) active cardiovascular disorder; 7) currently pregnant; 8) lifetime history of psychiatric disorder; 9) moderate to severe alcohol use disorder, or current substance use/dependence; 10) history of moderate to severe traumatic brain injury.

The sample was 64.3% female and 67.9% White/Caucasian. Ethnicity of our sample otherwise consisted of 14.3% Asian, 10.7% Multi-Ethnic or “Other,” and 3.6% African American. One individual elected not to report their ethnicity. Age ranged between 19-64 (M = 33.6 years; SD = 12.7).

## Materials

### Questionnaires and Task

#### Demographics

Participants were administered a brief demographics questionnaire regarding background information, including age, sex, and ethnicity.

#### State-Trait Anxiety Inventory (Spielberger et al., 1983)

The State-Trait Anxiety Inventory (STAI) is a 40 item self-report of trait and state anxiety, with 20 items assessing each trait and state symptoms. Higher scores on each sub-scale indicate higher levels of anxiety with scores ranging between 20 and 80 on each subscale. Internal consistency on the scale ranges between .86 and .96. In the current study, the internal consistency for the state version of the scale was α=.85 and for the trait version of the scale α=.77.

#### Beck Depression Inventory-II (Beck et al., 1996)

The Beck Depression Inventory-II (BDI-II) is a 21-item self-report inventory of depression symptoms, with higher scores indicating more severe levels of depression. Scores range between 0 and 63, and internal consistency ranges between .87 and .93. Internal consistency in the current study α=.77.

#### Active Visual Oddball P3 (Kappenman et al., 2021)

The Active Visual Oddball P3 task from the ERPCore task list developed by Kappenman and colleagues (2021) was used to elicit the P300 ERP. The letters A through E were presented (200ms; interstimulus interval of 1200-1400ms) in random order at an equal probability of 20% with one letter being designated as the target letter in each block of the task. In each block of the task, the target was presented at a rate of 20% and the non-targets were presented at a rate of 80%. Participants were asked to indicate the target and non-target using button presses. The task consists of 5 blocks consisting of 40 trials, for 200 total trials. Block order is randomized across participants. Full details on the task, as well as figures of sample trials and ERPs may be found in the ERPCore validation manuscript (Kappenman et al., 2021).

### TaVNS

The stimulation system consists of a commercially available, FDA-cleared constant current stimulator (Digitimer DS7AH, Digitimer Ltd., Hertfordshire, UK) with 1×3/8-inch hydrocolloid electrodes (Micro Neolead® ECG Electrodes, Neotech Products LLC, Valencia, CA). The active condition consisted of direct electrical stimulation delivered to the left cymba conchae (anode) and the tragus (cathode) based on prior work (Kreisberg et al., 2021). The left earlobe, used as a sham, has minimal ABVN innervation. All participants had electrodes attached for active and sham stimulation regardless of condition.

Stimulation current was delivered at 200% of each participant’s individual perceptual threshold (PT) and repeated for each stimulation condition (tragus and earlobe) (Badran et al., 2019). PT procedure entails single pulses of stimulation to the target area and obtaining verbal confirmation of stimulation perception while modulating the current intensity via parametric estimation by sequential testing (PEST) method (Badran et al., 2019; Taylor & Creelman, 1967). This allowed for determination of the minimum current intensity perceived at a specific target location and has been described in other studies by this group.

The sham stimulation condition utilized identical stimulation parameters as the active condition: 500 μs pulse width and 25 Hz frequency in 60 second on/off trains (Badran, Dowdle et al., 2018; Badran, Mithoefer et al., 2018). Active dose bins for each active dose duration group are presented in Table 1.

**Table 1.**
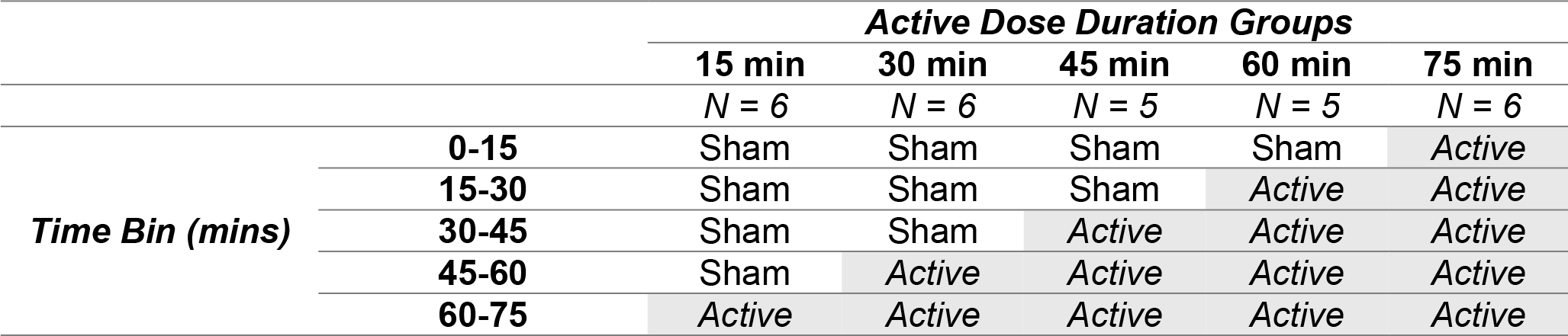
Dose duration bins for each active duration group.

|  |  | **Active Dose Duration Groups** |  |  |  |  |
| --- | --- | --- | --- | --- | --- | --- |
|  |  | **15 min** | **30 min** | **45 min** | **60 min** | **75 min** |
|  |  | *N* = 6 | *N* = 6 | *N* = 5 | *N* = 5 | *N* = 6 |
| **Time Bin (mins)** | **0-15** | Sham | Sham | Sham | Sham | Active |
|  | **15-30** | Sham | Sham | Sham | Active | Active |
|  | **30-45** | Sham | Sham | Active | Active | Active |
|  | **45-60** | Sham | Active | Active | Active | Active |
|  | **60-75** | Active | Active | Active | Active | Active |

Participants were blinded to which dose bin they were assigned, as was the primary investigator. Participants were set up for both active and sham taVNS across each dose condition to ensure blinding was maintained. A nonblinded coordinator switched the leads in the Digitimer output at the end of each dose bin depending on duration condition. In order to ensure blinding was maintained, the nonblinded coordinator unplugged and imitated lead switching at the end of each block, as the sound and movement at a particular time could lead to unblinding. All active dose bins were delivered contiguously and at the end of the dosing period, so cumulative effects could be assessed.

### Psychophysiological Recording and Processing

#### Electroencephalography

Continuous electroencephalography (EEG) and electrooculography (EOG) were recorded using a typical 32-channel montage actiCHamp amplifier (Brain Vision, Inc.) using Ag/AgCl electrodes mounted in an elastic cap in the standard 10/20 system. Data were referenced online to an electrical signal within the amplifier and then re-referenced to electrode Fz. Vertical EOG were measured using an electrode placed approximately 1 cm under the left eye in line with the pupil. In order to measure horizontal EOG, two other electrodes were placed near the outer canthi of the left and right eye. Data were sampled at 500 Hz.

EEG processing was completed using EEGLAB version 14 (Delorme & Makeig, 2004), ERPLAB version 7.0.0 (Lopez-Calderon & Luck, 2014). Data were average re-referenced and filtered offline using a .01-.30 Hz band-pass filter (Luck, 2014). To account for 25 Hz noise in the data attributable to taVNS, the ZapLine-plus plugin for EEG lab also was utilized. A notch filter was not used as the artifact was unstable during taVNS on/off trains. ZapLine-plus, instead, uses a moving spatial and spectral filter that can effectively remove temporally unstable frequency specific noise (Klug & Kloosterman, 2022). ERPLAB automated routines were used to detect and reject artifacts. Independent components analysis was applied to the EEG to detect and correct blinks and saccades. A 200ms window was moved through the data in 50ms increments to detect changes in the voltage due to artifact including flat lining, movement, or other noise lasting for the duration of the window. Trials with artifacts in the baseline or measurement window were excluded and participants whose data had more than 25% of trials rejected were not included in analyses (Luck, 2014). The P300 was measured at 350ms-450ms and baseline corrected (−200ms-0ms) and was utilized as an indicator of stimulus discrimination between rare and frequent stimuli and an indirect index of LC-NE activity.

#### Heart Rate

Heart rate was acquired via electrocardiogram (ECG) Biopac module. Three disposable pre-gelled Ag/AgCl electrodes were used: one each on the right and left forearms, and a third (ground) on the right ankle. Data were sampled at 1000 Hz. A band pass filter .5-35 Hz was applied to the data (Ruha et al., 2002). A template correlation function in Acknowledge was utilized to transform the data, and then a single-epoch HRV spectral analysis, which uses a modified Pan and Tompkins algorithm, was applied to the heart rate data collected across the Oddball task (Pan & Tompkins, 2007). This detects R-R intervals which were then visually checked and corrected, and then HRV spectral analysis was applied to the data epoch, which decomposes the heart rate into different frequency bands. The following frequency bands were extracted: low frequency (LF; 0.04 – 0.15 Hz), and high frequency (HF; 0.15 – 0.40). Sympathetic-vagal balance (SVB) also was computed (HF/LF ratio).

#### Procedures

After participants were screened and provided informed consent, they were sent self-report measures to complete remotely. Once at the lab, participants were provided with a pregnancy test, as the effects of taVNS on a fetus are currently unknown. Although no tests had positive results, participants were informed during consent that a positive test would result in their being compensated and released. Upon arrival to the lab, participants were provided with additional surveys, including the BDI-II, a self-report of their health behaviors (sleep, exercise, as well as caffeine, alcohol, and substance use), both typically and in the last 24 hours. These were administered in lab due to time sensitivity and for researchers to be able to respond to endorsement of suicidal ideation on the BDI-II.

Then electrodes were adhered for psychophysiology. Other recording devices were also set up for additional study procedures not presented in this pre-print. TaVNS set-up procedures including PT were completed. Participants were then administered a cognitive battery, which included the Oddball P300 (T1). Following administration of the tasks, taVNS was delivered for 60 minutes while innocuous nature films were played to occupy participants. During the final 10 minutes of taVNS, participants completed the cognitive battery a second time (T2). Following completion of these tasks, a taVNS tolerability and blinding questionnaire was completed, then participants were compensated for their time and released.

## Results

### Preliminary analyses

#### Psychological Measures

Descriptive statistics were run to evaluate demographics and relevant psychological variables to ensure that the sample was psychologically healthy. Scores indicated mild levels of trait anxiety (M = 29.3; SD = 5.8) and state anxiety (M = 26.2; SD = 5.7), as well as no/minimal past 2-week depression (M = 2.7; SD = 3.0).

#### Perceptual Threshold

In the preliminary analyses we also evaluate PT descriptives. Descriptive statistics are presented in Table 2. Dosing of the intensity of taVNS is still under development. Here, as we previously stated, 200% of each participant’s individual PT was utilized, consistent with prior work from our team (Badran et al., 2019). Other research groups may use the same stimulation intensity for all participants, though PT allows for maximization of tolerability for stimulation. Given this increases risk for a potential confound, data were evaluated for average PT within each dose group across active and sham sites and between dose groups. PTs were also evaluated in a repeated-measure Dose Group × Site ANOVA for interaction effects. Results indicated no main effect of Dose Group F(4,21)=.55, *p*=.70, no main effect of Site F(1,21)=1.22, *p*=.28, and no interaction effect F(4,21)=1.27, *p*=.31. Pairwise comparisons also were examined and revealed no spurious differences, all *p*s>.05. These data suggest that stimulation intensity did not significantly vary within and between active/sham locations or dose groups, which could otherwise impact interpretation of main results.

**Table 2.** Mean and standard deviation (mA) of the PT for each dose group at each of the active and sham locations.

|  |  | ***Dose Duration Group*** |  |  |  |  |  |
| --- | --- | --- | --- | --- | --- | --- | --- |
|  |  | **15** | **30** | **45** | **60** | **75** | **Total** |
| ***Active Tragus*** | **Mean** | 1.88 | 1.47 | 2.32 | 1.80 | 1.48 | 1.78 |
|  | **SD** | 0.58 | 0.76 | 1.10 | 0.00 | 0.89 | 0.77 |
| ***Sham Lobe*** | **Mean** | 2.20 | 1.40 | 1.76 | 2.52 | 2.32 | 2.02 |
|  | **SD** | 0.55 | 0.49 | 0.65 | 1.61 | 2.35 | 1.28 |

#### Descriptive Statistics for Primary Outcomes

Descriptive statistics for HRV and P300 data were computed for the baseline (T1) measurement and for the end of the dosing period (T2). Means and standard deviations for the HRV and P300 indices are presented in Table 3. Raw scores for HRV indices and P300 are shown in Figure 1 (difference scores presented in Figures 3 and 4). ERP waveforms and scalp maps are shown in Figure 2.

**Table 3.** Descriptive statistics for study time points.

| Dose Duration Group |  | 15 |  | 30 |  | 45 |  | 60 |  | 75 |  |
| --- | --- | --- | --- | --- | --- | --- | --- | --- | --- | --- | --- |
|  |  | *M* | *SD* | *M* | *SD* | *M* | *SD* | *M* | *SD* | *M* | *SD* |
| T1 | *Symp. Power* | 0.67 | 0.15 | 0.61 | 0.23 | 0.76 | 0.06 | 0.50 | 0.21 | 0.46 | 0.15 |
|  | *Vagal Power* | 0.33 | 0.15 | 0.39 | 0.23 | 0.24 | 0.06 | 0.50 | 0.21 | 0.54 | 0.15 |
|  | *SVB Power* | 2.79 | 2.27 | 2.42 | 2.04 | 3.40 | 1.17 | 1.48 | 1.52 | 0.99 | 0.65 |
|  | *P300* | 1.47 | 1.41 | 3.45 | 2.03 | 1.86 | 1.56 | 2.46 | 2.89 | 2.04 | 1.06 |
| T2 | *Symp. Power* | 0.70 | 0.23 | 0.62 | 0.26 | 0.74 | 0.06 | 0.45 | 0.25 | 0.45 | 0.24 |
|  | *Vagal Power* | 0.30 | 0.23 | 0.38 | 0.26 | 0.26 | 0.06 | 0.55 | 0.25 | 0.55 | 0.24 |
|  | *SVB Power* | 4.18 | 3.19 | 2.87 | 2.38 | 3.07 | .89 | 1.31 | 1.47 | 1.15 | .99 |
|  | *P300* | 2.36 | 2.06 | 3.95 | 2.96 | 1.71 | 2.51 | 2.19 | 2.37 | 2.07 | 1.29 |

**Figure 1.**
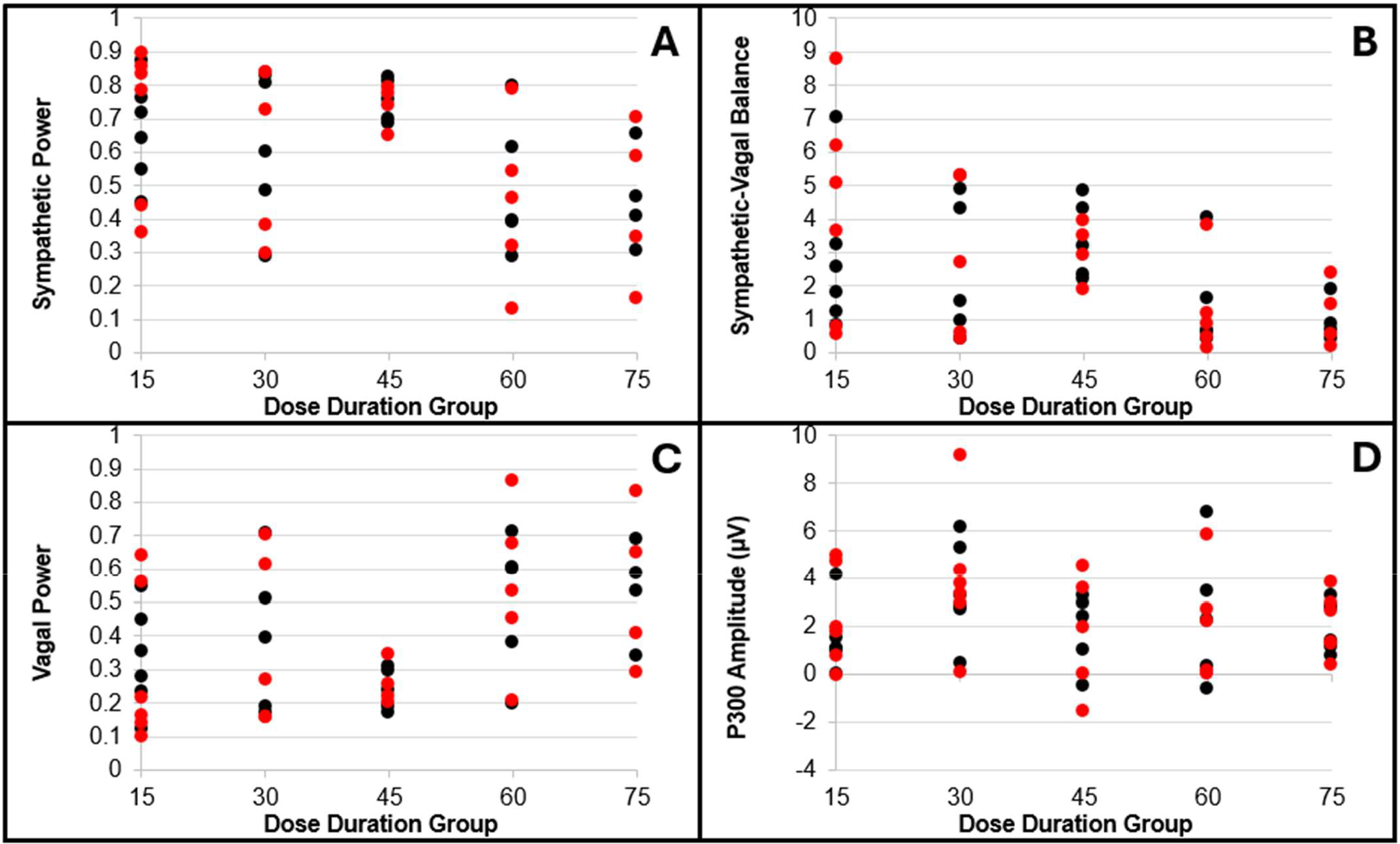
T1 and T2 scores for each outcome variable. Note. T1 is plotted in black and T2 is plotted in red.

**Figure 2.**
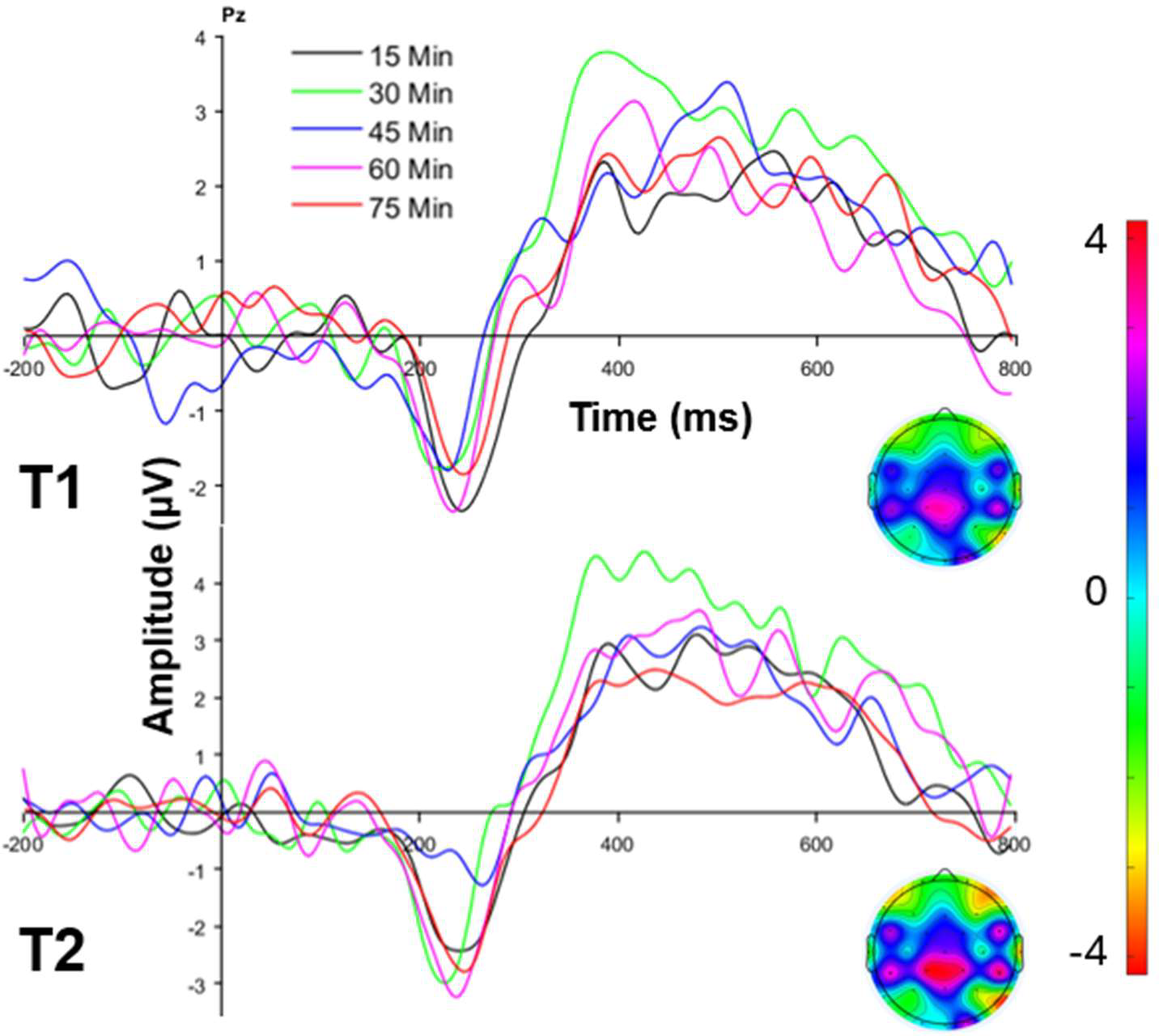
P300 waveforms and scalp maps for T1 and T2, plotted by dose duration group. Note. Scalp map amplitude is indicated via color, shown on the right bar red (negative) to magenta (positive). Individual P300 amplitude scores are plotted above in Figure 1, D. Here, they are presented at the group level.

**Figure 3.**
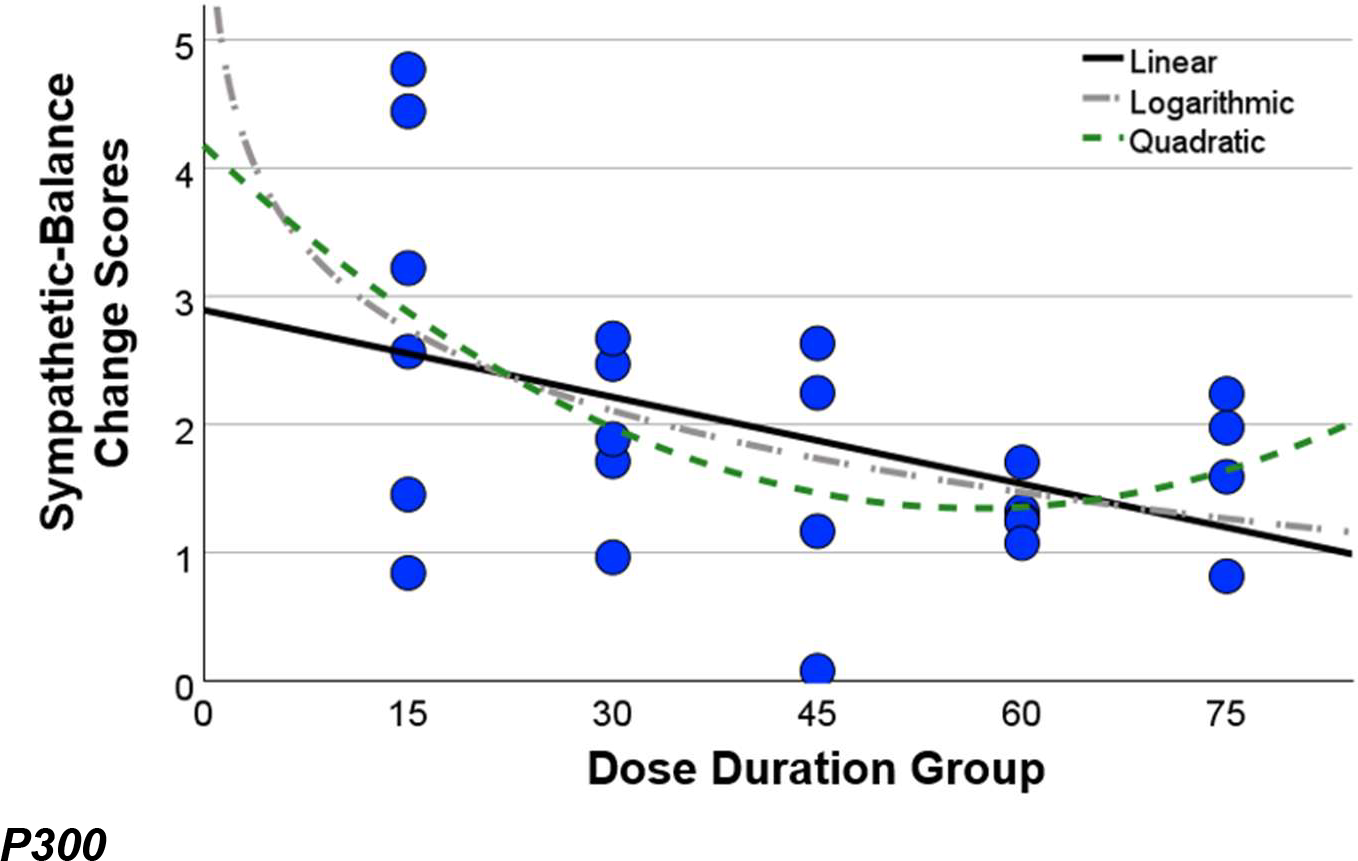
SVB change scores plotted by dose duration group, as well as linear, logarithmic, and quadratic curves.

**Figure 4.**
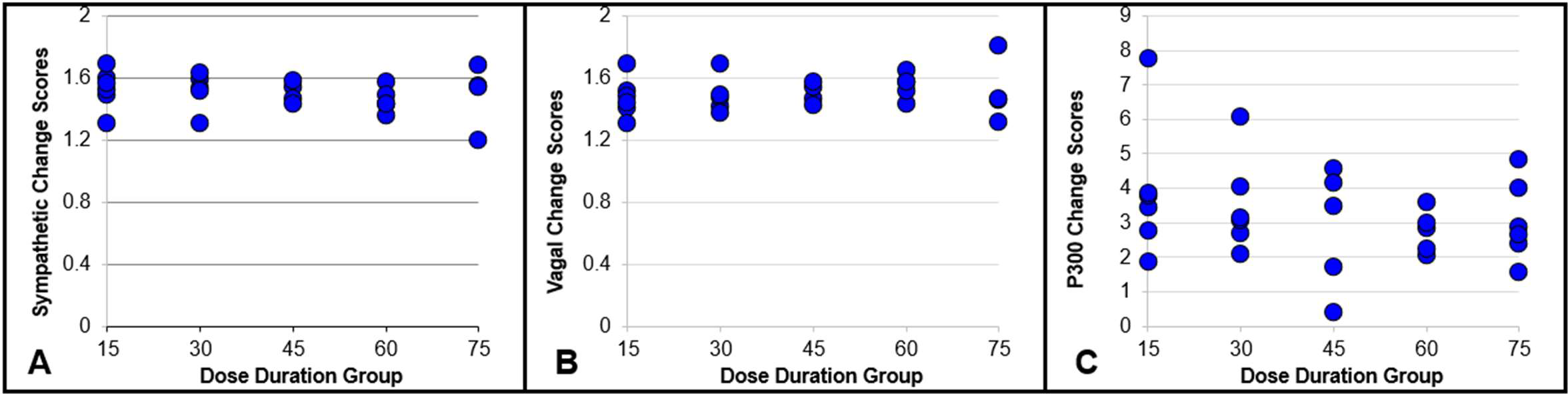
Change scores for non-significant outcomes (a) sympathetic, b) vagal, c) P300 change scores) by dose duration group.

### Primary Analyses

Following examination of raw T1 and T2 scores, change scores were computed for evaluation of curve estimation. Change scores were computed using a subtraction method and the indices of HRV and the P300 data were screened to ensure they met all assumptions for curve estimation and regression, including random distribution and normality of residuals. Because some measurements were negative, a constant was added to all change score outcomes to allow for evaluation of logarithmic model fit. Primary analyses consisted of the use of curve estimation regression models (linear, logarithmic, and quadratic) on indices of HRV and the P300 across active dose durations (15-75). For each model ANOVAs were calculated along with regression coefficients, R squared, standard error of the estimate and power.

#### Heart Rate Variability

Results indicated that for each index of HRV, there were no significant linear models (sympathetic or vagal) with the exception of SVB. The standardized coefficient for this effect, *B*=-0.45, SE=0.01, t(22)=-2.37, *p*=0.03, indicated that greater dose durations resulted in a greater reduction in SVB. See Table 4 for summary of results.

**Table 4.** ANOVA results, p values, R square and standard error of the estimate are presented for linear models.

|  | **ANOVA** | **p** | **R<sup>2</sup>** | **SE** |
| --- | --- | --- | --- | --- |
| **Sympathetic** | F(1,22)=0.73 | 0.40 | 0.03 | 0.12 |
| **Vagal** | F(1,22)=0.73 | 0.40 | 0.03 | 0.12 |
| **SVB** | F(1,22)=5.63 | 0.03* | 0.20 | 1.00 |

There was a significant logarithmic effect for SVB, but no other HRV indices (sympathetic or vagal; Table 5). The standardized coefficient for this effect, *B*=-0.51, SE=-0.33, t(22)=-2.75, *p*=-0.01, indicated that greater dose durations resulted in a greater reduction in SVB. Figure 2 demonstrates this effect, and the curve fit, indicating that at least 60 minutes of taVNS results in the greatest SVB reduction.

**Table 5.** ANOVA results, p values, R square and standard error of the estimate are presented for logarithmic models.

|  | **ANOVA** | **p** | **R<sup>2</sup>** | **SE** |
| --- | --- | --- | --- | --- |
| **Sympathetic** | F(1,22)=0.79 | 0.39 | 0.03 | 0.12 |
| **Vagal** | F(1,22)=0.79 | 0.39 | 0.03 | 0.12 |
| **SVB** | F(1,22)=7.09 | 0.01* | 0.26 | 0.97 |

There were no quadratic effects for any of the HRV indices, again, with the exception of SVB (Table 6). The standardized coefficients for the Dose effect, *B*=-2.00, SE=0.05, t(22)=-2.09, *p*=0.049 and the Dose effect**2, *B*=1.57, SE=0.001, t(22)=1.65, *p*=0.11 indicated that greater dose durations resulted in a greater reduction in SVB, though the curve of this model was not significant. Figure 3 displays SVB change score plots along with linear, logarithmic, and quadratic curves.

**Table 6.** ANOVA results, p values, R square and standard error of the estimate are presented for quadratic models.

|  | **ANOVA** | **p** | **R<sup>2</sup>** | **SE** |
| --- | --- | --- | --- | --- |
| **Sympathetic** | F(2,21)=0.41 | 0.67 | 0.04 | 0.12 |
| **Vagal** | F(2,21)=0.41 | 0.67 | 0.04 | 0.12 |
| **SVB** | F(2,21)=4.40 | 0.03* | 0.30 | 0.97 |

### P300

Results indicated that for the P300, there was no modulation in change scores by dose duration. See Table 7 for summary of results. P300 raw and scores are plotted in Figure 1, panel D, whereas change scores are plotted in Figure 4, panel C.

**Table 7.** ANOVA results, p values, R square and standard error of the estimate are presented for P300 ERP curve estimation models.

|  | ANOVA | *p* | R<sup>2</sup> | SE |
| --- | --- | --- | --- | --- |
| **Linear** | F(1,22)=0.81 | 0.38 | 0.04 | 1.52 |
| **Logarithmic** | F(1,22)=1.17 | 0.29 | 0.05 | 1.51 |
| **Quadratic** | F(2,21)=1.02 | 0.38 | 0.09 | 1.51 |

## Discussion

The current study was a sham controlled, blinded evaluation of the taVNS (15-75 active minutes) dose duration response of several psychophysiological responses in a sample of healthy controls. Here, we present HRV and ERP data from baseline to the end of the dosing period, as an initial test of the dose duration effect. Results partially supported our hypothesis. Data indicated that, while neither sympathetic nor vagal responses alone showed a dose duration response, greater taVNS durations (i.e., 60-75 mins) did result in decreased SVB – suggesting a particular impact on the ratio of sympathetic to vagal influence on the heart in the direction of increasing vagal predominance. Unlike HRV, the P300 ERP data, then did not show a dose duration dependent response. These data provide preliminary evidence for an autonomic response, but not a neural response during engagement of attention.

HRV data indicated that greater doses of taVNS resulted in decreased SVB akin to responses observed during relaxation breathing or meditation. Analyses, more specifically, indicated that there was a negative linear, logarithmic, and quadratic effect. However, the quadratic effect did not have a significant beta coefficient for the curve. This, along with comparable r-square indices and standard error of the mean, suggests that a logarithmic effect may be the better fit to the data. A lack of statistically significant asymptote to the data may indicate that even longer durations of taVNS may be warranted. Alternatively, visual inspection of the curve and data suggest that accruing a larger sample could lend greater power to the quadratic effect and the associated curve. In all, these data, so far, indicate that 60 to 75 minutes of taVNS are needed to adaptively decrease SVB, a dose duration that is much longer than is commonly used in the literature (Kim et al., 2022).

Observation of SVB scores alone does not help discriminate if taVNS results in an increase in vagal balance (parasympathetic response) or a decrease in sympathetic balance. Here, our sympathetic and vagal scores did not indicate modulation by taVNS dose duration, suggesting that there is a within subject, individualized response or a combinatorial effect on autonomic responses. In other words, an individual may have initially presented with a sympathetic preponderance and vagal tone may increase through taVNS, and another individual might present with low vagal tone and taVNS resulted in a sympathetic decreases to match this response more or less. However, these findings suggest the individual differences in preponderance may be less important and the end result is more adaptive. We are continuing to analyze skin conductance response data, which will help elucidate the effect of taVNS dose duration on sympathetic responses. In addition, future investigation with a larger sample at these higher doses could assist in parsing individualized and/or combinatorial effects. These findings could potentially support taVNS as a neuromodulation technique that could be applied based on individual autonomic responses. This could be beneficial in the context of certain spectrum-like psychiatric conditions (e.g., posttraumatic stress disorder) that present with either predominant hyperarousal or depressive/ affective blunting across individuals.

Unlike HRV, P300 data were not modulated by dose duration. The P300 is an ERP that is associated with attention broadly, and more specifically, stimulus discrimination, which has also been linked to LC-NE activation, a purported mechanism of taVNS (Warren et al., 2019). Whereas some previous research has demonstrated modulation of P300 via taVNS, our data are consistent with prior null findings (Burger et al., 2020; Warren et al., 2019; Warren et al., 2020). These results suggest there may be differential effects on gradual (autonomic or affective states) vs. instantaneous (electrocortical or event-related) processes. TaVNS may affect affective states while leaving event-related responding intact.

In addition, it is necessary to interpret the current P300 findings in the context of the ERPCORE Oddball task and the current healthy sample. The Oddball task is a relatively easy speeded response task, with no affective components. A more motivationally engaging task (e.g., fear learning, reward processing, affective go/no go) may help disentangle attentional vs. motivationally salient effects of stimulation. In addition, the P300 is an indicator of higher order stimulus discrimination, as opposed to orienting or attentional engagement. Planned analyses of steady-state visually evoked potentials collected in this study during passive viewing will provide additional evidence to help distinguish attention allocation vs. attention discrimination.

As mentioned, the current sample was healthy. This was a necessary approach for determining target engagement across multiple doses to ensure neural and autonomic changes were specific to taVNS rather than psychological or other symptoms. However, this strategy may come with limitations. Here, modulation of autonomic function via taVNS dose may be limited, given this output may be “optimized” in a healthy sample. While we did observe relaxation-like effects at higher doses, this observation may vary in a psychiatric sample. Specifically, individuals with a condition may move from maladaptive to normative responding. On the other hand, the oddball P300 may have a ceiling effect among healthy individuals, as the ERPCore oddball is not highly demanding or motivationally engaging. TaVNS may engage the P300 where there are deficits among a sample with psychiatric symptoms, indicating engagement of LC-NE activity, though our sample did not have observable deficits.

Altogether, these findings suggest that an important next step is to evaluate these effects in a sample with a psychiatric condition which may have greater variability in the purported taVNS targets. This would allow for replication of the possible optimal dose (60-75 mins) and evaluate shared mechanisms of taVNS and psychiatric disorders. Furthermore, as with many brain stimulation candidates, taVNS may affect multiple psychiatrically relevant processes (e.g., fear extinction) and outputs (e.g., heart rate variability). These effects may be convergent, but often are discordant, as is the case here. Therefore, future investigation should continue to evaluate multi-modal responses, as well as multiple relevant processes.

The current data add to a growing body of evidence that a) demonstrates that taVNS adaptively modulates vagal tone, b) has narrowed the parameter space of taVNS, c) suggests that others are significantly underdosing taVNS, which could contribute to accumulating null effects, and d) adds to the case for inclusion of multiple outputs when investigating novel brain stimulation candidates. To our knowledge this is the first ever dose duration finding study for taVNS and future investigations are needed to continue to narrow the parameter space and replicate these findings in a larger sample with psychiatric symptoms.

## Data Availability

All data produced in the present study are available upon reasonable request to the authors

